# Phase 2 Randomized Trial of an AS03 Adjuvanted Plant-Based Virus-Like Particle Vaccine for Covid-19 in Healthy Adults, Older Adults and Adults with Comorbidities

**DOI:** 10.1101/2021.05.14.21257248

**Authors:** Philipe Gobeil, Stéphane Pillet, Iohann Boulay, Annie Séguin, Alexander Makarkov, Gretchen Heizer, Kapil Bhutada, Asif Mahmood, Nathalie Charland, Sonia Trépanier, Karen Hager, Julia Jiang-Wright, Judith Atkins, Matthew P. Cheng, Donald C. Vinh, Philippe Boutet, François Roman, Robbert Van Der Most, Maria Angeles Ceregido, Marc Dionne, Guy Tellier, Jean-Sébastien Gauthier, Brandon Essink, Michael Libman, Jason Haffizulla, André Fréchette, Marc-André D’Aoust, Nathalie Landry, Brian J. Ward

## Abstract

The rapid spread of SARS-CoV-2 continues to impact humanity on a global scale with rising total morbidity and mortality. Despite the development of several effective vaccines, new products are needed to supply ongoing demand and the needs of specific populations. We report herein a pre-specified interim analysis of the phase 2 portion of an ongoing Phase 2/3, randomized, placebo-controlled trial of a coronavirus virus-like particle (CoVLP) vaccine candidate produced in plants that displays the SARS-CoV-2 spike glycoprotein adjuvanted with AS03 (NCT04636697). A total of 753 subjects were recruited between 25 November 2020 and 24 March 2021 into three groups: Healthy Adults (18-64 years: N=306), Older Adults (≥ 65 years: N=282) and Adults with Comorbidities (≥18 years: N=165) and randomized 5:1 to receive two intramuscular doses of either vaccine CoVLP (3.75 μg/dose + AS03) or placebo 21 days apart. This report presents safety, tolerability and immunogenicity data collected up to 21 days after the second dose. The immune outcomes presented include neutralizing antibody (NAb) titres and cellular (IFN-γ and IL-4 ELISpot) responses. In this study, CoVLP+AS03 was well-tolerated and adverse events (AE) after each dose were generally mild to moderate and transient. Solicited AEs in Older Adults and Adults with Comorbidities were generally less frequent than in Healthy Adults. CoVLP+AS03 induced seroconversion in >35% of subjects in each group after the first dose and in ∼98% of subjects 21 days after the second dose. In all treatment groups, NAb levels were ∼10-fold higher than those in a panel of convalescent sera. A significant minority (∼20%) of subjects had evidence of a pre-existing IFN-γ response to the S protein and almost all subjects in all groups (>88%) had detectable cellular responses (IFN-γ, IL-4 or both) at 21 days after the second dose. A Th1-biased response was most evident after the first dose and was still present after dose two. These data demonstrated that CoVLP+AS03 will likely be well-tolerated and highly immunogenic in adults ≥18 years of age with and without comorbidities.

## Introduction

Following a cluster of pneumonia cases in the city of Wuhan in Hubei province of China in December 2019 ^1^, a novel coronavirus (Severe Acute Respiratory Syndrome Coronavirus 2 [SARS CoV-2]) was identified as the causative agent. The disease was subsequently named ‘coronavirus disease 2019’, or COVID-19 ^1,2^. The rapid international spread of COVID-19 prompted the World Health Organization (WHO) to declare a pandemic in March 2020 ^3^. As of October 10^th^, 2021 there has been more than 236 million cases of COVID-19 and ∼4.8 million deaths ^4^. This public health emergency sparked a remarkable global effort to develop vaccines using a wide range of traditional and novel platforms including messenger ribonucleic acid (mRNA), deoxyribonucleic acid (DNA), inactivated virus, live viral vectors, recombinant proteins, peptides, or virus-like particles (VLPs) ^5,6^. As of October 10^th^, 2021, >90 of these vaccines have entered clinical testing and at least 20 of them have been authorized for use in at least one country. Despite these successes, there remains an urgent global need to identify additional safe and effective vaccines.

While it is now clear that humoral immunity is highly correlated with protection ^7^, both innate and cellular immunity ^8-11^ likely also contribute to protection against SARS-CoV-2 infection. Passive antibody transfer has proven protective in both non-human primate animal models and in the early treatment of some patients ^12-14^. Correlates of protection have recently been proposed for both binding and neutralizing antibodies ^7,15,16^. In parallel, a role for cell-mediated immunity has been suggested for viral clearance and prevention of serious disease, as well as for long-term immunity ^13,17-19^. Optimally, SARS-CoV-2 vaccines would provide a well-coordinated response engaging multiple elements of the immune system.

The vaccine candidate developed by Medicago, a Coronavirus-like particle (CoVLP), is a self-assembling VLP that displays trimers of recombinant modified S protein of SARS-CoV-2 (ancestral variant) embedded in a lipid envelope. The VLPs are produced in a plant (*Nicotiana benthamiana*) and closely resemble the native structure of SARS-CoV-2 ^20,21^. The VLPs are administered with an oil-in-water adjuvant: Adjuvant System 03 manufactured by GSK (AS03, hereafter CoVLP+AS03) ^22^. AS03 initiates a transient innate immune response at the injection site and draining lymph node in animal models ^23,24^ and in human peripheral blood ^9,25,26^ that can potentiate and shape both antibody and T-cell responses ^27-29^. AS03 has been used in the licensed pandemic A/H1N1pdm09 influenza vaccines Arepanrix H1N1 (in Canada) and Pandemrix (in Europe), of which 90 million doses have been administered worldwide, as well as in other licensed (Q Pan H5N1 in the USA) or candidate vaccines ^30^. In Medicago’s Phase 1 study, AS03 significantly enhanced both cellular and humoral responses to CoVLP and the vaccine showed a favorable safety profile ^21^.

Herein we report results of the Phase 2 portion of an ongoing Phase 2/3 randomized, placebo-controlled study conducted at multiple sites in Canada and the USA. The Phase 2 portion of this study was designed to confirm ^21^ the chosen formulation and dosing regimen (2 doses of 3.75 μg CoVLP adjuvanted with AS03 given 21 days apart). This report includes responses in healthy adults aged 18-64 (“Healthy Adults”), in adults ≥65 years of age (“Older Adults”) and in adults with comorbidities (“Adults with Comorbidities”). Assessment of the safety and efficacy of CoVLP+AS03 in all three adult populations is currently ongoing in the global, Phase 3 portion of the study.

## Results

### Demographic and baseline clinical characteristics

Subjects were screened for SARS-CoV-2 antibodies using a commercial ELISA that targets the nucleocapsid protein (although both seronegative and seropositive subjects were enrolled) and randomized 5:1 to receive CoVLP+AS03 or placebo (saline). Participant demographics are presented in Table 1 and subject disposition is presented in Figure 1. The mean ages of Healthy Adults were 43.7 years and 42.4 years in vaccinated and placebo groups, respectively. The mean ages of Older Adults were 71.1 years and 71.7 years in vaccinated and placebo groups, respectively. Finally in Adults with Comorbidities, the mean ages were 56.6 and 57.1 in the vaccinated and placebo groups respectively. In all groups, subjects were mostly White or Caucasian (96.7% in Healthy Adults, 98.6% in Older Adults, and 81.8% in Adults with Comorbidities). A minority of subjects in the three populations self-identified as Hispanic or Latino (2.9% in Healthy Adults, 1.8% in Older Adults, and 14.5% in Adults with Comorbidities), Asian (2.0%, 1.1%, and 0%) or Black or African American (1.0%, 0.0%, and 17.6%). The Phase 3 portion of the Phase 2/3 study is being conducted in six countries globally and includes a more diverse population than that recruited in Phase 2.

**Table 1:** Summary of demographics and baseline characteristics (NCT04636697)

|  | Healthy Adults |  | Older Adults |  | Adults with Comorbidities |  |
| --- | --- | --- | --- | --- | --- | --- |
|  | CoVLP<br>3.75 µg<br>+AS03 | Placebo | CoVLP<br>3.75 µg<br>+AS03 | Placebo | CoVLP<br>3.75 µg<br>+AS03 | Placebo |
| Subjects | 259 | 47 | 235 | 47 | 138 | 27 |
| Sex, n (%) |  |  |  |  |  |  |
| Male | 112<br>(43.2) | 23 (48.9) | 110<br>(46.8) | 19 (40.4) | 77 (55.8) | 16 (59.3) |
| Female | 147<br>(56.8) | 24 (51.1) | 125<br>(53.2) | 28 (59.6) | 61 (44.2) | 11 (40.7) |
| Race, n (%) |  |  |  |  |  |  |
| American Indian or<br>Alaska Native | 0 | 0 | 1 (0.4) | 0 | 1 (0.7) | 0 |
| Asian | 5 (1.9) | 1 (2.1) | 3 (1.3) | 0 | 0 | 0 |
| Black or African<br>American | 2 (0.8) | 1 (2.1) | 0 | 0 | 23 (16.7) | 6 (22.2) |
| Native Hawaiian or<br>Other Pacific Islander | 0 | 0 | 0 | 0 | 0 | 0 |
| White or Caucasian | 251<br>(96.9) | 45 (95.7) | 231<br>(98.3) | 47 (100) | 114<br>(82.6) | 21 (77.8) |
| Not Reported | 1 (0.4) | 0 | 0 | 0 | 0 | 0 |
| Age at the time of<br>informed consent (years) |  |  |  |  |  |  |
| Mean (SD) | 43.7<br>(13.00) | 42.4<br>(13.29) | 71.1<br>(5.14) | 71.7<br>(4.87) | 56.6<br>(13.75) | 57.1<br>(12.69) |
| Median | 45.0 | 45.0 | 70.0 | 72.0 | 58.0 | 58.0 |
| Min, Max | 18, 64 | 18, 64 | 65, 88 | 65, 87 | 23, 82 | 24, 81 |
| SARS-CoV-2 at Baseline,<br>n (%) |  |  |  |  |  |  |
| Positive | 8 (3.1) | 0 | 5 (2.1) | 1 (2.1) | 13 (9.4) | 4 (14.8) |
| Negative | 247<br>(95.4) | 46 (97.9) | 225<br>(95.7) | 46 (97.9) | 122<br>(88.4) | 23 (85.2) |

Max, maximum; Min, minimum; n, number of subjects in categories; SD, standard deviation.

**Figure 1.**
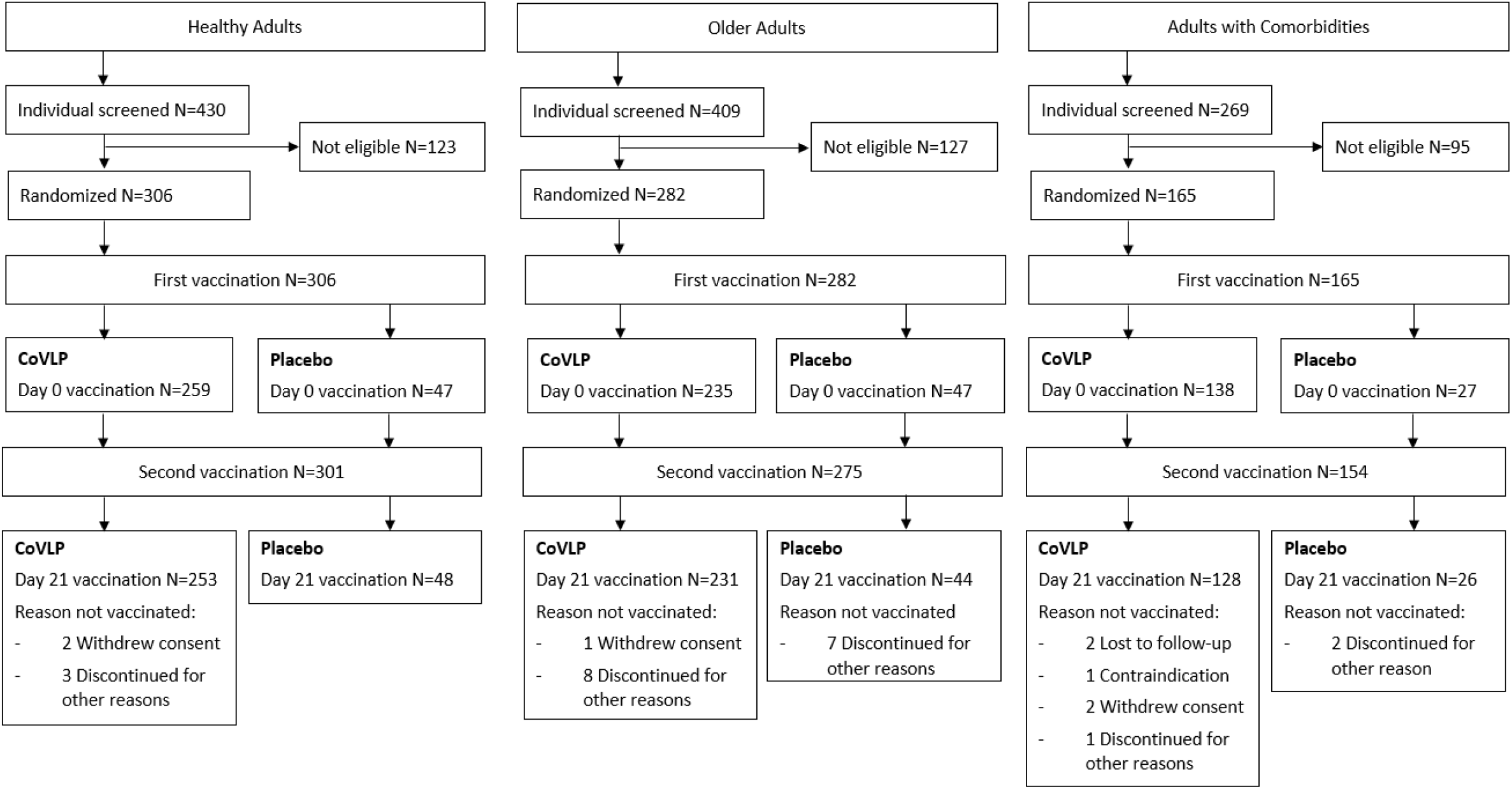
Trial Profile – Subject Disposition. Enrollment and follow-up of study participants vaccinated with CoVLP with AS03 or placebo after the first and second dose administration. For both Adults and Older Adults, one subject was mis-dosed for the second dose administration and received a placebo in error. One individual in the Adults with Comorbidities group that was initially seropositive at D0 did not receive an injection. For more details of subject disposition, see Table 1.

Three hundred and six Healthy Adults, 282 Older Adults, and 165 Adults with Comorbidities were enrolled in the study between 25 November 2020 and 24 March 2021. Of the 753 subjects who received a first injection (placebo or vaccine), 730 (96.7%) also received a second injection.

### Safety

Safety and tolerability data after the first and second doses are provided for 306 and 301 subjects in the Healthy Adults, 282 and 275 subjects in the Older Adults, and 165 and 154 subjects in the Adults with Comorbidity groups respectively. Overall, the vaccine was well-tolerated in all populations, with a slightly milder reactogenicity profile in the Older Adults (apart from slightly greater rates of erythema) and Adults with Comorbidities.

Reactogenicity is illustrated in Figure 2 for solicited A) local and B) systemic AE. Frequency of at least one solicited AE in vaccinated individuals increased after the second dose relative to the first dose in Healthy Adults and Older Adults although Older Adults generally reported fewer solicited AEs: 88.4 and 94.5% of Healthy Adults reported at least one AE after the first and second doses respectively compared to 66.8 and 83.5% of the Older Adults. The pattern of increased reactivity at second dose was not observed in Adults with Comorbidities where the frequency of solicited AE was 71.0% and 64.8% after first or second doses respectively.

**Figure 2.**
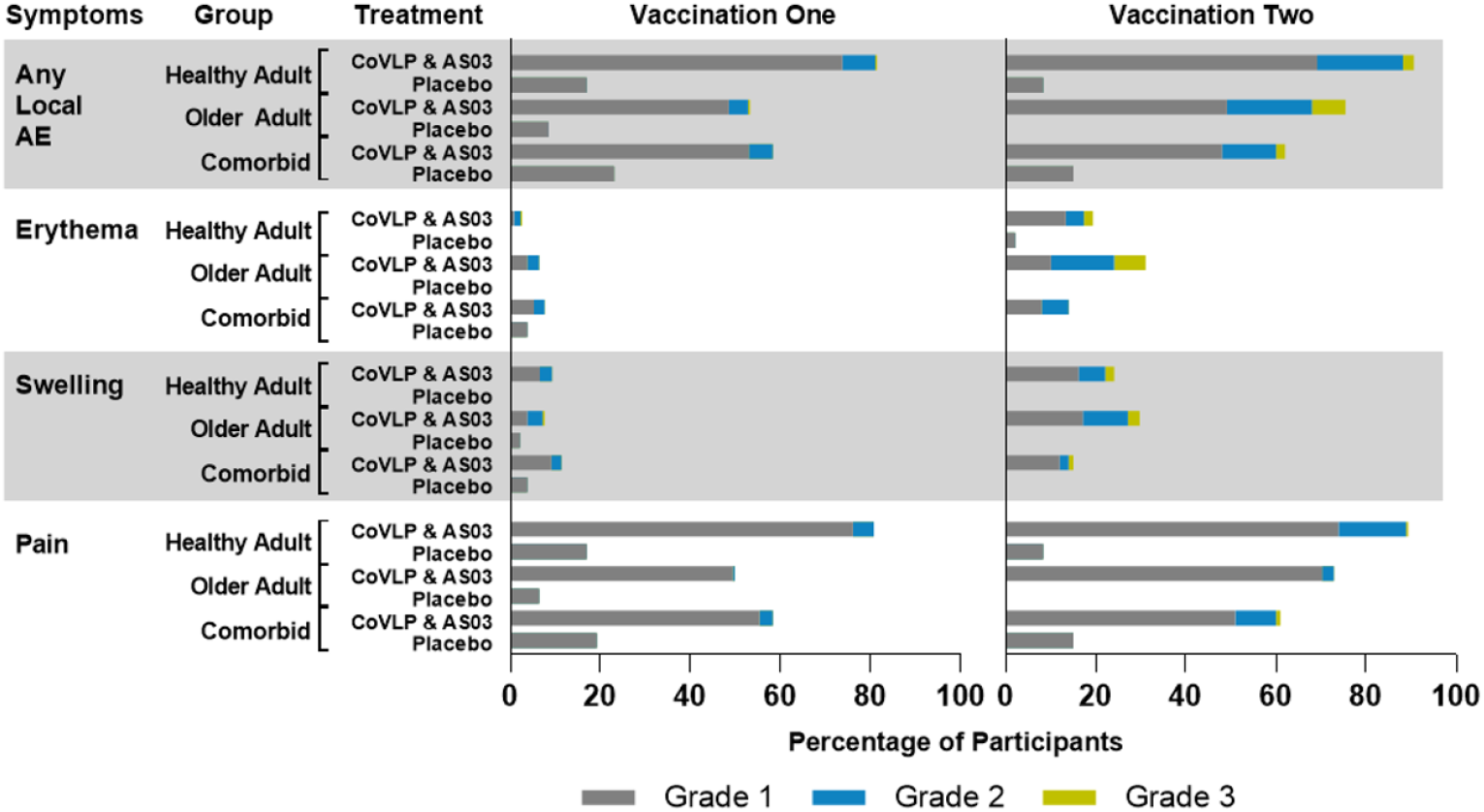
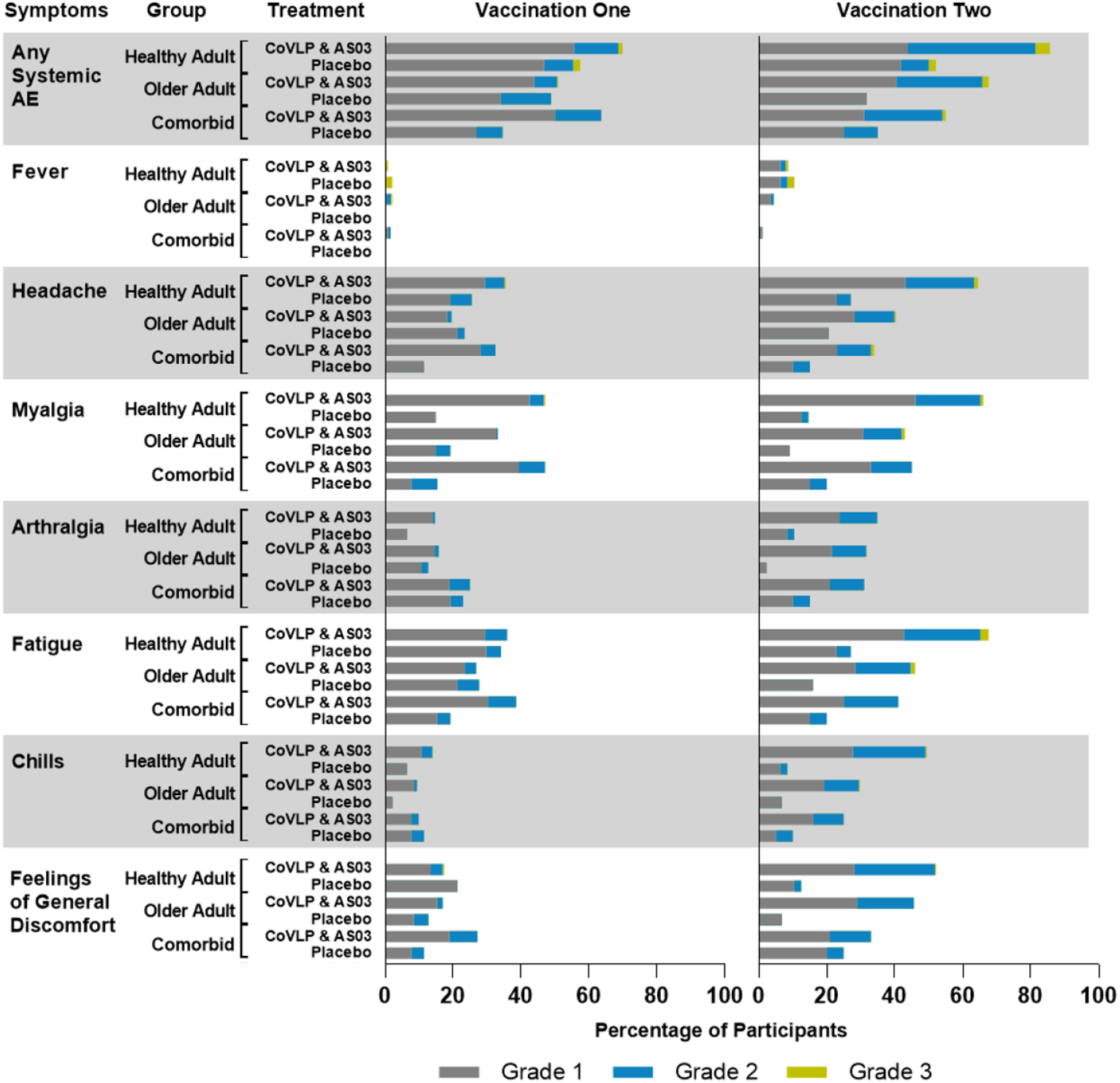
Solicited Local and Systemic Adverse Events 7 Days After the First or Second Vaccine Candidate Dose. Subjects were monitored for solicited local (panel A) and systemic (panel B) AEs from the time of vaccination through 7 days after administration of each dose. No Grade 4 (potentially life-threatening) events were reported. Participants who reported no adverse events (AEs) make up the remainder of the 100% calculation (not shown). If any of the solicited AEs persisted beyond 7 days after each vaccination (when applicable), it was recorded as an unsolicited AE. Fever was defined as oral temperature ≥38.0°C.

After the first dose, the most frequently reported solicited local AE was injection site pain (80.7% of Healthy Adults, 49.8% of Older Adults, 57.2% of Adults with Comorbidities) while the most frequently reported solicited systemic AE were fatigue (35.9% of Healthy Adults, 26.8% of Older Adults, 38.4% of Adults with Comorbidities), myalgia (46.7% of Healthy Adults, 33.2% of Older Adults, 46.4% of Adults with Comorbidities) and headache (35.5% of Healthy Adults, 19.6% of Older Adults, 32.6% of Adults with Comorbidities). After the second dose, pain at the injection site was the most frequently reported solicited local AE (89.3% of Healthy Adults, 73.6% of Older Adults, 60.9% of Adults with Comorbidities) while fatigue (67.6% of Healthy Adults, 46.8% of Older Adults, 43.8% of Adults with Comorbidities), myalgia (66.0% of Healthy Adults, 43.3% of Older Adults, 48.4% of Adults with Comorbidities), and headache (64.0% of Healthy Adults, 40.7% of Older Adults, 33.6% of Adults with Comorbidities) were the most frequently reported systemic AEs.

In all populations, the majority of at least one solicited local or systemic AEs were mild (Grade 1) or moderate (Grade 2) in severity and transient, typically resolving within 24 hours to 3 days. Grade 3 solicited AEs were reported by 1.5% and 6.3% of Healthy Adults after the first and second doses respectively, while in Older Adults 0.9% and 8.7%, and in Adults with Comorbidities 0% and 3.1% of participants experienced grade 3 AEs after the first and second doses respectively. A greater frequency of grade 3 erythema was observed post second dose in Older Adults (6.9%) relative to Healthy Adults (2.0%). No Grade 4 AE was reported by any subject. No clinically significant laboratory abnormalities, related serious AEs, or cases of Vaccine-Associated Enhanced Disease (VAED), anaphylaxis, or potential immune-mediated disorders monitored as Adverse Events of Special Interest (AESI) and meeting the protocol defined definition have been reported at the data cut-off of the phase 2 report. Only one non-serious case of grade 3 skin allergic type reaction was reported. This subject had a personal and family history of allergies to various materials. As of October 10^th^, 2021, two pregnancies have been reported 2.5 and 7 months after the second dose.

### Immunogenicity: Antibody Response

Ancestral strain-specific pseudovirion neutralizing antibody (NAb) responses are illustrated in Figure 3 and shown in Supplemental Table 1. Relative to both pre-vaccine sera (Baseline) and placebo controls, significant increases were observed in GMTs in all three cohorts at 21 days (D21) after the first dose with further significant increases 21 days after the second dose (D42).

**Figure 3.**
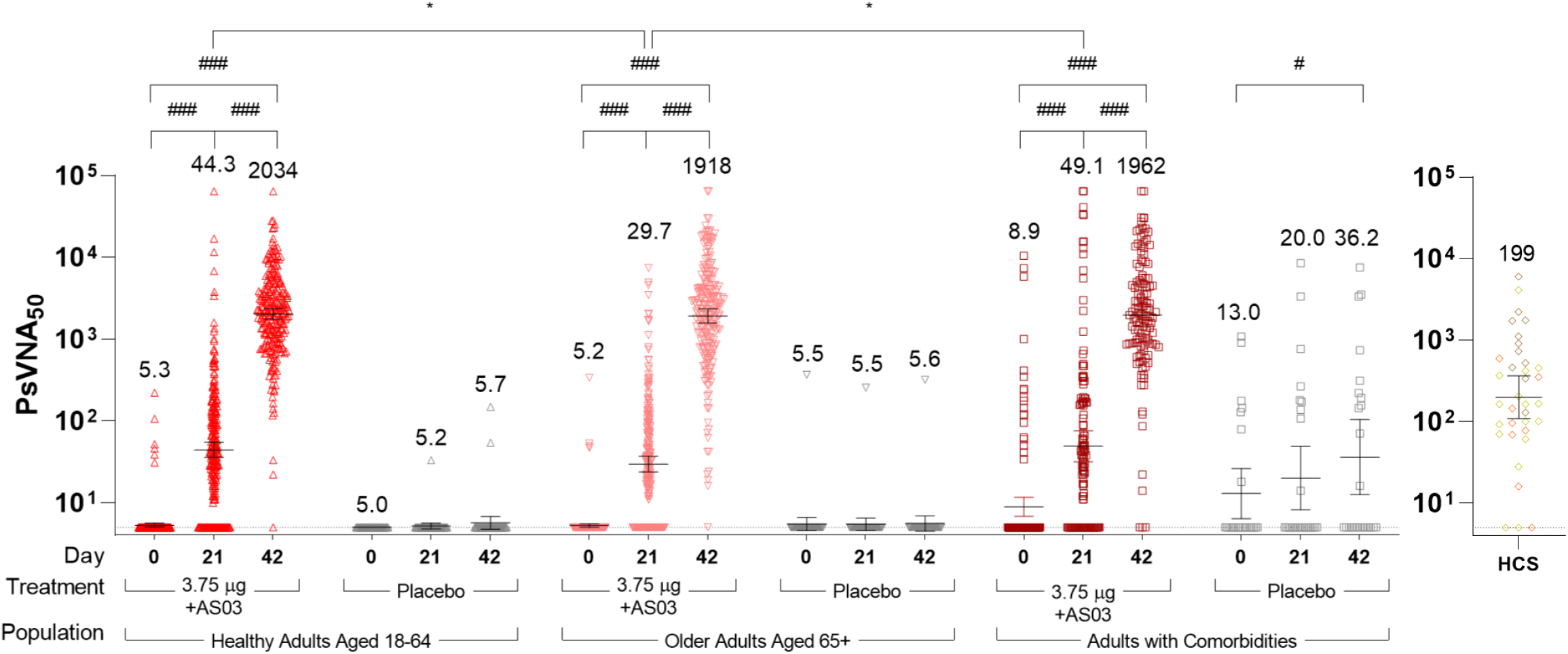
Humoral Response in Healthy Adults, Older Adults, and Adults with Comorbidities. Pseudovirion neutralization titers at baseline (Day 0) and 21 days after the first (Day 21) or the second dose (Day 42) of CoVLP +AS03 or placebo. Bars and numerical values indicate geometric means and error bars indicate 95% CI. Significant differences between Days 0, 21 and 42 are indicated by # (^###^*p*<0.001; paired T-test of log-transformed values, SAS). Significant differences across study populations at D21 or D42 are indicated by *(\**p*<0.05; one-way ANOVA on log-transformed data for GMT, SAS).

**Figure 4.**
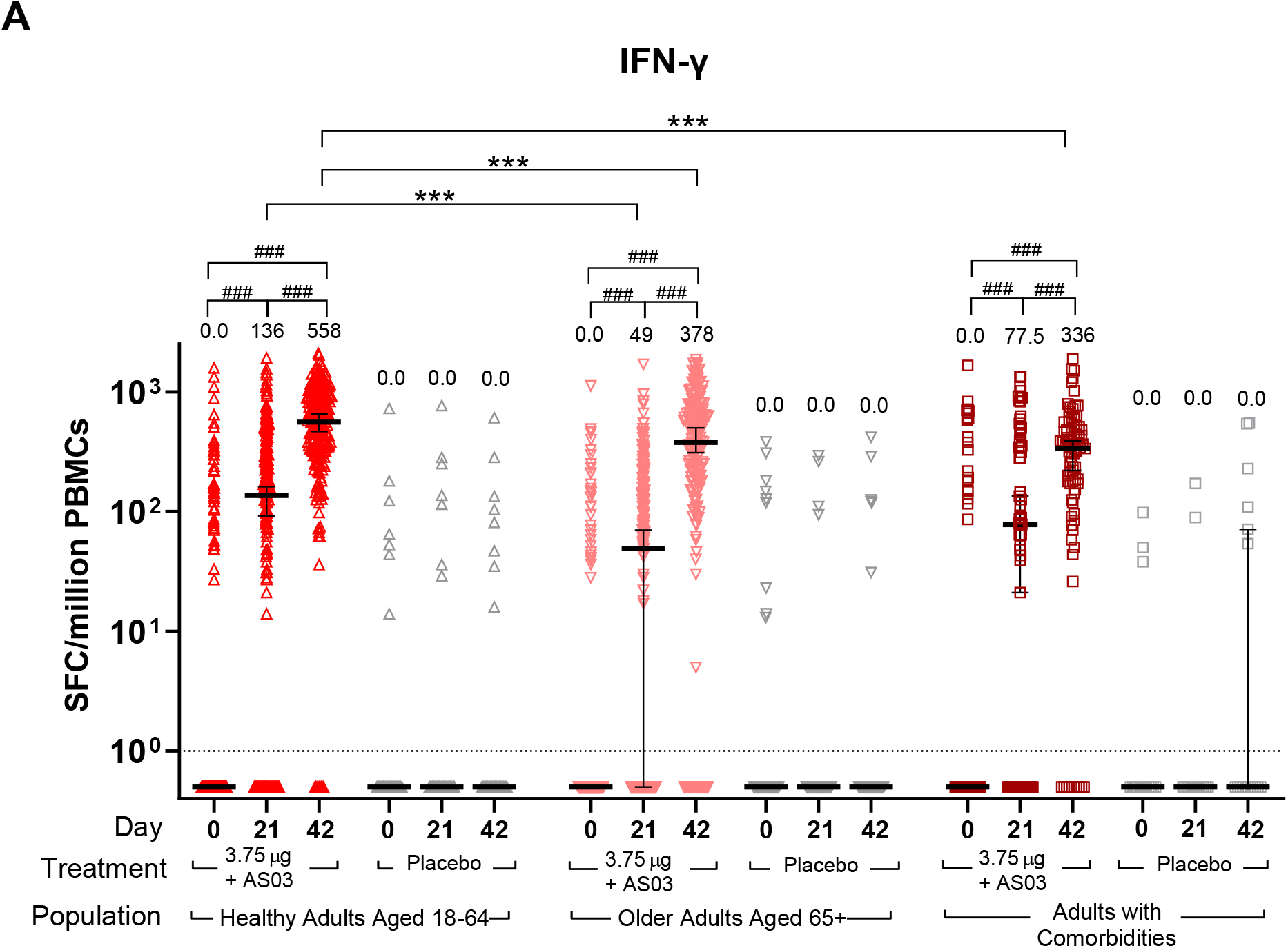
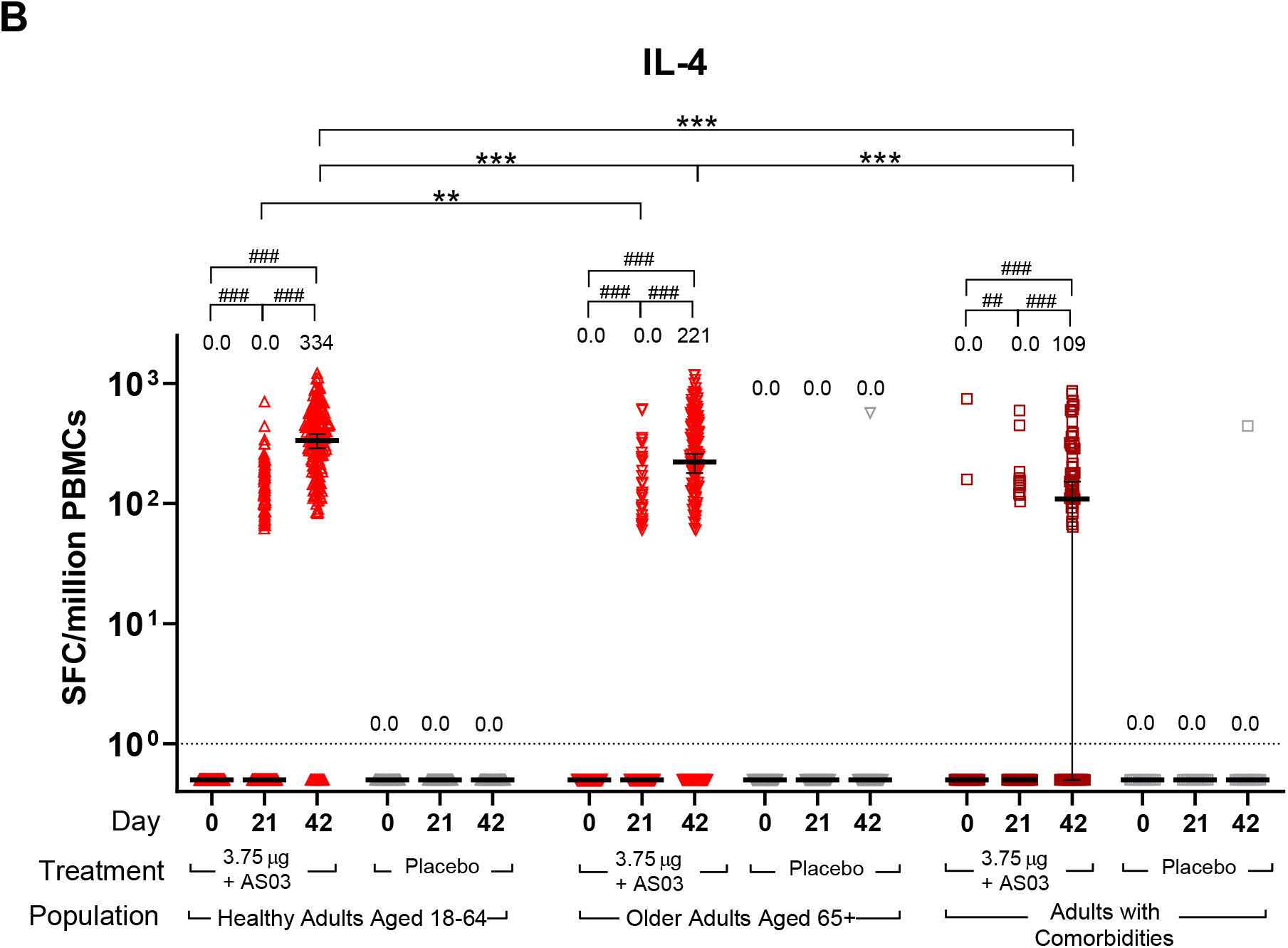
Cellular Immune Response in Healthy Adults, Older Adults, and Adults with Comorbidities. IFN-γ and IL-4 ELISpot Responses. IFN-γ (A) and IL-4 (B) spot forming cell (SFC) counts at baseline (Day 0), 21 days after the first immunization (Day 21) and 21 days after the second immunization (Day 42) with CoVLP (3.75 μg) adjuvanted with AS03 or placebo are represented. Bars indicate median (± 95% CI). Results of statistical analysis are represented for relevant comparisons. Significant differences between time-points for each vaccine regimen are indicated by # (## *p*<0.01, ###*p*<0.001; Wilcoxon signed rank test, SAS). Significant differences between study populations at D21 and D42 are indicated by * (\**p*<0.05, \*\*\**p*<0.001; Kruskal-Wallis test, GraphPad Prism v9.0.1). Both IFN-γ and IL-4 responses in subjects injected with placebo were significantly lower (not represented, Wilcoxon rank sum test, SAS) than subjects who received the adjuvanted CoVLP at both Day 21 and Day 42 regardless of the population cohort. An arbitrary value of 0.5 SPC was assigned to samples with undetectable values to allow for representation on a logarithmic scale.

A single dose of CoVLP+AS03 induced a four-fold rise in NAb (seroconversion) in a slightly larger proportion of the Healthy Adults (51.3%) relative to Older Adults (38.4%, *p*<0.01), an effect also reflected in the D21 GMTs (44.3, in Healthy Adults and 29.7, in Older Adults, *p*<0.05). There was no significant difference in seroconversion between Healthy Adults and Adults with Comorbidities (44.2%) or in D21 GMTs (49.1). The differences in seroconversion rates and GMTs between the Healthy Adults and Older Adults disappeared with the second dose of CoVLP+AS03. The GMTs at D42 were 2034 for Healthy Adults and 1918 in Older Adults. The seroconversion rates were 99.2% in Healthy Adults and 97.7% in Older Adults. Between Healthy Adults and Adults with Comorbidities at D42, there were no statistical differences in GMT (1962 in Adults with Comorbidities) although a difference in seroconversion (95.8% in Adults with Comorbidities, *p*<0.05) was observed.

Consistent with observations from the Phase 1 clinical trial, the NAb titers elicited by CoVLP+AS03 at D42 were approximately 10-fold higher than those observed in a panel of convalescent sera (10.2x in Healthy Adults, 9.6x in Older Adults, and 9.9x in Adults with Comorbidities).

To assist in standardizing the NAb results, the WHO reference standard 20/136, pooled plasma from individuals with particularly high titers ^31^, was included in the pseudovirion NAb assay yielding a reference titer of 1872. Expressing the GMT results in International Units (IU/mL) after the first and second doses respectively (by dividing by 1.872), the Healthy Adults in our Phase 2 study had NAb values of 23.7 and 1087 IU/mL (at D21 and D42), the Older Adults had values of 15.9 and 1025 IU/mL, and the Adults with Comorbidities had values of 26.2 and 1048 IU/mL. Using this methodology, HCS had a value of 106 IU/ml.

Prior to vaccination, 31 (4.12%) of the subjects the Phase 2 portion of the study were seropositive at D0, 25 of whom received CoVLP+AS03: 8 Healthy Adults (GMT at D0 32.3, 95% confidence interval (95CI): 8.6, 121.9), 6 Older Adults (GMT 46.0, 95CI: 7.2, 294.1) and 16 Adults with Comorbidities (GMT 141.6, 95CI: 39.4, 509.7). The Nab response was robust in these subjects at both D21 with GMT of 3078 (95CI: 397.8, 23810), 1762 (95CI: 174.4, 17787), and 1466 (95CI: 181.5, 11842) and at D42 with GMT of 7426 (95CI: 2620, 21048), 6918 (95CI: 1189, 40259), and 7042 (95CI: 2630, 18858), in Healthy Adults, Older Adults and Adults with Comorbidities respectively. Day 21 titres were significantly higher in these seropositive subjects than the initially seronegative subjects in all groups (*p*<0.001, 4171, 31.33 respectively).

Overall, two doses of CoVLP +AS03 induced strong and comparable NAb responses in all groups and a single dose of COVLP+AS03 was sufficient to generate a robust response in seropositive subjects. Although small differences between Healthy Adults and Older Adults were observed at D21 after the first dose, these differences disappeared with the second dose.

### Immunogenicity: Cell Mediated Response

Vaccination with CoVLP+AS03 induced a significant (*p*<0.001) increase of the IFN-γ response at D21 in all groups. This marker of a Th1-type response was further increased (*p*<0.001) after a second dose at D42 in the three populations while no effect was observed in placebo recipients (Figure 3A). Healthy Adults vaccinated with CoVLP+AS03 had a higher IFN-γ response compared to Adults with Comorbidities at D42 (*p*<0.001) and to Older Adults at both D21 and D42 (both *p*<0.001). The IFN-γ response at D0 significantly correlated (Spearman correlation test, *p*<0.001) with detection of NAb at baseline and this positive correlation was maintained after both the first and the second doses in vaccinated subjects. As observed during the Phase 1 CoVLP clinical trial (NCT0445004) ^21^, a substantial minority of individuals (21%, 17% and 27% in Healthy Adults, Older Adults and Adults with Comorbidities respectively) had pre-existing (D0) IFN-γ ELISpot responses to S protein. The proportion of subjects with a detectable IFN-γ response was 69% of the Healthy Adults, 54% of Older Adults and 63% of the Adults with Comorbidities after one dose (D21) and reached 96% (Healthy Adults) and 88% (Older Adults and Adults with Comorbidities) after the second dose.

Also consistent with the results from the CoVLP Phase 1 clinical trial, with the exception of two Adults with Comorbidities, no subject had a measurable IL-4 response pre-vaccination. Vaccination with CoVLP+AS03 significantly (*p*<0.01) increased the IL-4 responses after one dose and was further significantly (*p*<0.001) increased after a second dose in the three populations while no effect was observed in placebo groups (Figure 3B). After two doses, Adults with Comorbidities had a significantly (*p*<0.001) lower IL-4 response than either Healthy Adults or Older adults. The IL-4 response was also significantly lower in Older Adults relative to Healthy Adults after one (*p*<0.01) or two (*p*<0.001) doses. While a limited portion of subjects responding to CoVLP+AS03 elicited an IL-4 response after one dose (33%, 17% and 18% in Healthy Adults, Older Adults and Adults with Comorbidities respectively), the proportion of responders increased to 92% (Healthy Adults), 79% (Older Adults) and 62% (Adults with Comorbidities) after the second dose of CoVLP+AS03. The number of Th1-type (IFN-γ) responding cells was consistently 1.7 to 3.0-fold greater than the Th2-type (IL-4) cells in all groups 21 days after the second dose.

## Discussion

The Phase 2 portion of the ongoing Phase 2/3 study of CoVLP+AS03 was designed to confirm the selection of CoVLP dose and adjuvant identified in the Phase 1 trial and to assess the performance of the chosen formulation in Healthy Adults, Older Adults, and Adults with Comorbidities. The primary outcomes for the Phase 2 portion of this study focused on short-term (up to D42) safety and tolerability of CoVLP+AS03 and the ability of this novel vaccine candidate to induce both NAb and cellular responses to SARS-CoV-2 spike protein.

Reactogenicity in Healthy Adults in the Phase 2 study confirmed the profile observed in the much smaller Phase 1 study in 18–55-year-old adults that received the CoVLP (3.75μg) + AS03 formulation (n=20) ^21^. Local reactogenicity was primarily characterized by injection site pain in most subjects while systemic reactogenicity was primarily characterized by myalgia, fatigue, and/or headache in approximately 70% of the study participants. Relative to the first dose, there was a consistent trend towards increased local and systemic reactogenicity after the second dose in Healthy and Older Adults and towards increased local reactogenicity in Adults with Comorbidities; almost all reported AEs were mild-to-moderate and transient in nature. Although only the 3.75 μg dose level of CoVLP was used in the Phase 2 study, data from the Phase 1 trial that included both unadjuvanted and AS03-adjuvanted groups at three CoVLP dose levels (3.75, 7.5 and 15 μg/dose) showed that local and systemic reactogenicity was largely attributable to AS03, as would be expected from its use with other antigens ^25,32,33^. As expected ^26^ and has been observed for mRNA, adenovirus vector, inactivated virus, and recombinant protein-based COVID-19 vaccines ^34-41^ slightly reduced reactogenicity was observed in Older Adults. Consistent with reports using other COVID-19 vaccine in Adults with Comorbidities ^42^, the reactogenicity in this population was not increased relative to that seen in Healthy Adults. Overall, no new safety concerns were raised in this study, suggesting that CoVLP+AS03 would be well-tolerated in adults ≥18 years of age.

Both the humoral and cellular immune responses seen in the Phase 2 study confirmed the robust immunogenicity results documented in the 18-55 year old Healthy Adults during the Phase 1 study ^21^ as well as extending these observations to Older Adults and Adults with Comorbidities. After two doses of CoVLP+AS03, seroconversion was observed in ∼98% of the subjects and NAb titers were ∼10x higher than those found in convalescent sera. Although the NAb response after the first dose of CoVLP+AS03 was slightly reduced in the Older Adult population relative to the Healthy Adults, the differences between the two cohorts disappeared after the second dose. This observation of reduced immunogenicity in Older Adults after the first dose is consistent with the generally reduced capacity of older individuals to respond to vaccination and with findings with other SARS-CoV-2 vaccines ^41,43,44^. The decreased ability of even healthy older individuals to mount strong immune responses after vaccination is likely multi-factorial including a general decline in immune function (ie: immunosenescence) and chronic low-level inflammation (so-called ‘inflammaging’) ^45^. Consistent with the observations that adjuvants can enhance vaccine-induced responses in older individuals ^46^, these results suggest that two-doses of CoVLP+AS03 can overcome the age-associated limitations for NAb production at least.

The number of subjects in the current study with pre-existing NAb titers to SARS-CoV-2 was low (n=31, 4.12%) but vaccination of these individuals with CoVLP+AS03 nonetheless induced a substantial increase in titers, suggesting that CoVLP+AS03 can significantly boost a pre-existing memory response. This observation, consistent with other SARS-COV-2 vaccines ^47-49^ provides strong support for vaccinating both infection naïve and previously infected individuals.

Overall, the NAb titers induced by CoVLP+AS03 in all populations compared favourably with a panel of convalescent serum/plasma. While this method can be used to draw broad comparisons between studies ^7,15^, there is a growing consensus that this approach has limitations for comparing responses between groups and between trials with different vaccines. For this reason, we included the WHO reference standard 20/136, pooled antibodies from recovered COVID-19 patients with very high NAb response in multiple laboratory studies ^50^ in our serological analysis so that the performance of CoVLP+AS03 could be assessed relative to other vaccines. This analysis confirmed the induction of very high NAb titres (>1000 IU/mL) by CoVLP+AS03 in all three groups studied. In this study the mean standardized value for the HCS was 106.3 IU/mL, a noticeably lower value than the 1000 IU/mL of the 20/136 WHO reference pool, a difference attributable to the selection of high titer individuals in the pooled WHO reference standard relative to the more diverse titers observed across the mild, moderate and severely ill individuals from which the convalescent sera in this study are drawn.

Although attention on vaccine-induced immune responses for SARS-CoV-2 has focused primarily on antibody production, there is compelling evidence that cellular responses contribute to both recovery from infection and long-term immunity ^13,51^. Despite this growing consensus, the optimal vaccine-induced cellular response is not yet fully understood ^52^. In the current study, the ‘balanced’ Th_1_ (IFN-γ) and Th_2_ (IL-4) cellular response induced by CoVLP+AS03 was, like the NAb response, entirely consistent with the Phase 1 results ^21^. Although an IFN-γ dominated response was observed after the first dose in the three populations, the strong Th1-bias evolved to include a substantial IL-4 induction after the second dose. Even though the IL-4 response increased significantly after the second dose, spot forming unit (SFU) counts for IFN-γ remained approximately 1.7 to 3-fold higher than those for IL-4. The limitation of using only these two ELISpot assays to assess the pattern of cellular immunity generated by CoVLP+AS03 is acknowledged.

While the possibility of VAED was initially a point of concern in COVID-19 vaccine development ^53,54^, there has since been no evidence of disease enhancement in animal models, clinical trials or real-world evidence reported to date. Specifically, there has been no suggestion that vaccine-induced Th_2_ responses are associated with VAED ^52^. On the contrary, it is possible that the balanced Th1/Th_2_-response induced by CoVLP+AS03 including strong IL-4 production, contributed significantly to the high NAb titers observed in vaccinated subjects via its role in T helper follicular cell involvement, optimal germinal center formation, and B-cell maturation ^18,55-57^. Such Th_2_-driven effects may also contribute to longevity of SARS-CoV-2 specific memory B-cell responses and longer-term immune response ^58^.

Although inducing a robust cellular immune response in Older Adults, the impact of age on both IFN-γ and IL-4 responses suggested that at least some aspects of the aged immune system cannot be overcome with multiple doses of an AS03-adjuvanted vaccine. Given the clear age-related differences in both the clinical manifestations of COVID-19 and the immune response generated by SARS-CoV-2 infection ^59^, it is not surprising that vaccine-induced responses might also differ between younger and older individuals ^60^. Indeed, similar age-related differences in immune responses have been reported for several of the SARS-CoV-2 vaccines in development or in use ^37,61^ and age-related differences in vaccine efficacy with some of the deployed vaccines have been reported ^62-64^. We also observed a significant reduction of the cellular immune response in Adults with Comorbidities relative to Healthy Adults even after two doses of CoVLP+AS03. Besides age, the spectra of comorbidities exhibited by the subjects in this study (detailed in Methods) does not allow speculation regarding specific mechanisms of action to explain this reduction as some of the implicated conditions can potentially promote pro-inflammatory responses while others or their associated treatments can be immunosuppressive. Given the high NAb titers induced by CoVLP+AS03 and the strong cellular response across all populations after two doses, it is unclear whether or not the *in vitro* differences in cellular responses will result in clinically relevant differences in protection. Such theoretical differences may only become apparent as the Phase 3 portion of the study progresses and/or with time, should CoVLP+AS03 be licensed.

This study as presented has several limitations. Whether or not the NAb induced by CoVLP+AS03 have activity against variants of concern is of obvious interest and these studies are underway. In addition, the limited demographic diversity of the Phase 2 study participants is acknowledged; this is primarily a reflection of the demographics of the study sites where the majority of these participants were recruited. Relative to both these considerations, the ongoing global Phase 3 efficacy portion of the trial is being conducted on three continents and is expected to provide efficacy results relative to multiple variants and in a much more diverse population.

In conclusion, this report of the Phase 2 portion of our ongoing Phase 2/3 study of CoVLP+AS03 provides evidence that this formulation is likely to be well-tolerated and highly immunogenic in adults ≥ 18 years of age. The Phase 3 portion of the study will determine if these results are confirmed in a larger population. Compared to either a panel of convalescent serum/plasma or the WHO standard serum reagent (20/136), the NAb response induced by CoVLP+AS03 was among the highest reported for any SARS-CoV-2 vaccine. Across the broad age range of study participants (18 – 88 years of age), almost all mounted either a strong NAb response, a Th1/Th2-pattern cellular response, or both, following two doses of CoVLP+AS03.

## Data Availability

Medicago Inc. is committed to providing access to anonymized data collected during the trial that underlie the results reported in this article, at the end of the clinical trial, which is currently scheduled to be 1 year after the last participant is enrolled, unless granted an extension. Medicago Inc. will collaborate with its partners (GlaxoSmithKline, Belgium) on such requests before disclosure. Proposals should be directed to. To gain access, data requestors will need to sign a data access agreement and access will be granted for non-commercial research purposes only. The following publicly-available databases were accessed to complete this work: GISAID database (https://www.gisaid.org/) and Genbank (https://www.ncbi.nlm.nih.gov/genbank/).

https://www.ncbi.nlm.nih.gov/genbank/

## Author Contributions

All authors contributed significantly to the submitted work. BJ Ward, N Landry, A Seguin contributed to all aspects of the clinical study from conception to completion. P Gobeil, S Pillet, I Boulay, A Makarkov, G Heizer, K Bhutada, A Mahmood, N Charland, S Trépanier, K Hager, J Jiang-Wright and J Atkins contributed to design and execution of the study as well as analysis and presentation of the data. P Boutet, F Roman, R Van Der Most and MA Ceregido contributed to analysis and presentation of data. MP Cheng and DC Vinh provided access to critical reagents and consulted on study design and execution. M Dionne, G Teller, JS Gauthier, B Essink, M Libman, J Haffizulla and A Frechette were critically involved in the execution of the clinical trial. All authors contributed to critical review of the data and the writing of the manuscript. All Medicago authors had full access to the data. BJW, MAD and NC made the decision to submit the manuscript.

## Acknowledgements

The authors acknowledge all the volunteers who participated in the study, the site investigators and their staff who conducted the studies with a high degree of professionalism as well as all the Medicago employees and contractors involved in the study. The authors also acknowledge Karyne Framand for her support identifying appropriate references.

## Methods

### CoVLP Vaccine Candidate and Adjuvant

The CoVLP vaccine candidate has previously been described in detail ^21^. Briefly, full-length spike protein from SARS-CoV-2 (strain hCoV-19/USA/CA2/2020) incorporating the modifications R667G, R668S, R670S, K971P, and V972P is expressed in *Nicotiana benthamiana* by transient transfection, resulting in spontaneous trimer formation and VLP assembly and budding. The VLPs are purified and shipped to the vaccination site where it is mixed with AS03 adjuvant prior to injection.

AS03 adjuvant, an oil-in-water emulsion containing DL-α-tocopherol (11.69 mg/dose) and squalene (10.86 mg/dose), was supplied by GlaxoSmithKline.

### Vaccine Preparation and Injection

CoVLP was available in single-dose vials (0.30 mL) at 15 μg/mL and stored at 2-8°C until shortly before use. The AS03 adjuvant was supplied in multi-dose vials (10 doses/vial). Immediately prior to use 0.3 mL of CoVLP and 0.3 mL of AS03 were mixed gently 1:1 volume:volume in the CoVLP vial and a 0.5 mL withdrawn for injection. All injections were administered intramuscularly in the deltoid using a 23g needle of appropriate length based on body mass index (BMI). The first and second doses were administered contralaterally when possible.

### Study Design

The phase 2 portion of the study is a randomized, observer-blinded, placebo-controlled study with male and female subjects. The study was approved by a central Institutional Review Board/ Research Ethics Review Board as well as the Health Products and Food Branch of Health Canada and was carried out in accordance with the Declaration of Helsinki and the principles of Good Clinical Practices. Written informed consent was obtained from all study participants prior to any study procedure. Subjects were offered modest compensation for their participation in this study (ie: time off work, displacement costs).

Subjects were screened up to 14 days in advance of the first vaccine administration and must have demonstrated a satisfactory baseline medical assessment by history, general physical examination, hematologic, biochemic, and serologic analysis. Although a test for SARS-CoV-2 antibodies was performed at screening using a commercial ELISA that targets the nucleocapsid (N) protein (ElecSys: Roche Diagnostics), both seronegative and seropositive subjects were enrolled.

For Healthy Adults, subjects had to be 18-64 years of age. For Older Adults, subjects had to be 65 years of age or older and to be non-institutionalized (eg, not living in rehabilitation centers or old-age homes; living in an elderly community was acceptable). For Adults with Comorbidities, most frequent comorbidities were appetite and general nutritional disorders (obesity), allergic conditions, vascular hypertensive disorders, lipid metabolism disorders, glucose metabolism disorders (diabetes mellitus) and joint disorders.

For both Healthy Adults and Older Adults, subjects must have been in good general health with no clinically relevant abnormalities that could jeopardize subject safety or interfere with study assessments, as determined by medical history, physical examination, and vital signs, and have had a body mass index less than 30 kg/m^2^. Adults with Comorbidities included subjects with one or more comorbid conditions that puts them at higher risk for severe COVID-19 such as obesity, hypertension, type 1 or type 2 diabetes, chronic obstructive pulmonary disease (COPD), cardiovascular diseases, chronic kidney diseases, or a compromised immune system (eg, treatment-controlled HIV infection, organ transplant recipients, or patients receiving cancer chemotherapy). Female subjects of childbearing potential must have had a negative pregnancy test result at screening and vaccination and used a highly effective method of contraception for one month prior to vaccination and at least one month after the last study vaccination. Exclusion criteria for Healthy Adults and Older Adults included i) any significant acute or chronic, uncontrolled medical or neuropsychiatric illness, ii) any chronic medical condition associated with elevated risk of severe outcome of COVID-19, iii) any confirmed or suspected current immunosuppressive condition or immunodeficiency, including cancer, HIV, hepatitis B or C infection, iv) current autoimmune disease, v) administration of any medication or treatment that could alter the vaccine immune response. In all three study populations exclusion criteria also included vi) administration of any vaccine within 14 days prior to vaccination or planned administration of any vaccine up to Day 28 of the study, vii) administration of any other SARS-CoV-2 / COVID-19, or other experimental coronavirus vaccine at any time prior to or during the study, viii) history of virologically-confirmed COVID-19, ix) rash, dermatological condition, tattoos, muscle mass, or any other abnormalities at injection site that could interfere with injection site reaction rating, x) use of prophylactic medications (eg, antihistamines [H1 receptor antagonists], nonsteroidal anti-inflammatory drugs [NSAIDs], systemic and topical glucocorticoids, non-opioid and opioid analgesics) within 24 hours prior to the vaccination to prevent or pre-empt symptoms due to vaccination, xi) history of a serious allergic response to any of the constituents of CoVLP, including AS03, xii) history of documented anaphylactic reaction to plants or plant components (including tobacco, fruits and nuts), xiii) personal or family (first-degree relatives) history of narcolepsy, xiv) history of Guillain-Barré Syndrome. Sentinel subjects (10 in each group) were first enrolled in Older Adults and Adults with Comorbidities groups, and unblinded safety data after each dose were reviewed by the IDMC. Enrollment into the Phase 2 portion of the study was closed on 25 March 2021.

The participants and the personnel collecting the safety information and working in testing laboratories remained blinded to treatment allocation. On D0, D21 and D42, serum and peripheral blood mononuclear cells (PBMC) were processed for immune outcomes. All safety information was collected, and all laboratory procedures were carried out by study staff blinded to treatment allocation. There were no major Protocol changes during the conduct of this study prior to the preparation of the current manuscript.

### Primary and Secondary Objectives

The primary objectives of the Phase 2 portion of the study were to assess safety and tolerability and immunogenicity to CoVLP+AS03 at 0, 21, and 42 days post vaccination compared to placebo in Healthy Adults, Older Adults, and Adults with Comorbidities.

Primary safety outcomes were the occurrence(s) of i) immediate AEs within 30 minutes after each vaccination; ii) solicited local and systemic AEs up to 7 days after each vaccination; iii) unsolicited AEs, serious AEs (SAEs), AEs leading to withdrawal, AESIs, and deaths up to 21 days after each vaccination; iv) normal and abnormal urine, and hematological and biochemical values.

Primary immunogenicity outcomes were i) NAb titers measured using a pseudovirion neutralization assays and ii) IFN-γ and IL-4 ELISpot responses at 21 days after each dose of vaccine.

A secondary safety outcome was the occurrence(s) of SAEs, AEs leading to withdrawal, AESIs, and deaths from 22 days after the last vaccination up to the end of the study. Secondary immunogenicity outcomes were immune responses measured at days 128, 201 and 386 post-vaccination.

The safety and immunogenicity data collected at later timepoints in this ongoing study will be released once study follow-up has been completed.

### Safety Assessments

Both passive (diary) and active monitoring of safety signals were performed for the first 42 days of the study and will be continued throughout the study. Active monitoring included telephone contacts with subjects one and eight days (D1, D8) after each vaccination as well as a site visit on D3 after vaccination. Solicited AEs were assessed by the subjects as Grade 1 to 4 (mild, moderate, severe, or potentially life-threatening) according to criteria previously described ^21^. Per protocol, all solicited events (local, systemic) are considered related events. Unsolicited AEs, and AEs leading to subject withdrawal were collected up to 21 days after each vaccination. Based on IDMC recommendation, the following event(s) could pause or halt the study for further review and assessment of the event(s): i) Any death, ii) Any vaccine-related SAE; iii) Any life-threatening (Grade 4) vaccine-related AE; iv) If 5 % or more of subjects in a single treatment group, experienced the same or similar listed event(s) that could not be clearly attributed to another cause: v) A severe (Grade 3 or higher) vaccine-related AE; vi) A severe (Grade 3 or higher) vaccine-related vital sign(s) abnormality; vii) A severe (Grade 3 or higher) vaccine-related clinical laboratory abnormality.

Subjects will return to the Investigator site on Days 128, 201, and 386 for safety follow-ups and immunogenicity assessments.

Safety signals for VAED, hypersensitivity reactions, and potential immune-mediated diseases were monitored as AESI as previously described ^21^.

### SARS-CoV-2 Pseudovirion Immunogenicity-Neutralization Assay, Convalescent Sera/Plasma and WHO Reference Standard

Full details of the pseudovirion neutralization assay (Nexelis, Quebec, Canada) have previously been described ^21^. Briefly, the assay is based on a genetically modified Vesicular Stomatitis Virus (VSV) from which the glycoprotein G was removed, and a luciferase reporter introduced. The modified VSV vector expresses full length SARS-CoV-2 S glycoprotein (NXL137-1 in POG2 containing 2019-nCOV Wuhan-Hu-1; Genebank: MN908947) from which the last nineteen amino acids of the cytoplasmic tail were removed (rVSVΔG-Luc-Spike ΔCT). Pseudovirions are mixed with vaccinee sera and the degree of neutralization quantified using human ACE-2 expressing VERO cells and reduction in luciferase-based luminescence. For each sample, the neutralizing titer was defined as the reciprocal dilution corresponding to the 50% neutralization (NT_50_), when compared to the pseudoparticle control. Samples below cut-off were attributed a value of 5 (half the minimum required dilution).

Results were compared to sera/plasma from COVID-19 convalescent patients. These were collected from a total of 35 individuals with confirmed diagnosis. Time between the onset of the symptoms and sample collection varied between 27 and 105 days. Four sera samples were collected by Solomon Park (Burien, WA, USA) and 20 sera samples by Sanguine BioSciences (Sherman Oaks, CA, USA); all were from non-hospitalized individuals. Eleven plasma samples were collected from previously hospitalized patients at McGill University Health Centre. Disease severity was ranked as mild (COVID-19 symptoms without shortness of breath), moderate (shortness of breath reported), and severe (hospitalized). These samples were analyzed in parallel of clinical study samples, using the assays described above. Demographic characteristics were previously described ^21^.

To facilitate the comparability of results across different trials, the WHO International Standard for anti-SARS-CoV-2 immunoglobulin (human; NIBSC code: 20/136) was established to allow conversion of neutralization assay titers into international units (IU/mL). This standard consists of pooled plasma obtained from eleven individuals recovered from SARS-CoV-2 infection and with high NAb titers. Upon multiple assessments using this validated PNA assay, a conversion factor of 1.872 was established. Hence, the antibody titers presented throughout this manuscript can be expressed as IU/mL by dividing the NT50 by this factor.

### Immunogenicity-Interferon-γ and Interleukin-4 ELISpot

PBMC samples from study subjects were analyzed by IFN*-*γ or IL-4 ELISpot (Caprion, Quebec, Canada) using a pool of 15-mer peptides with 11aa overlaps from SARS-CoV-2 S protein (USA-CA2/2020, Genbank: MN994468.1, Genscript, purity >90%). Full details of the methodology are detailed elsewhere ^21^.

### Analysis Populations and Statistical Analysis Plan

Randomization was managed by Syneos Health with Medicago oversight using Medidata Rave RTSM interactive randomization tool (2021.2.0, Medidata, USA). Statistical analysis and data presentation was conducted using SAS (SAS Institute, North Carolina) and Prism (GraphPad Software, San Diego).

The sample size of 753 subjects made it possible to perform the initial evaluation of the vaccine immunogenicity and detect major differences in rates of AEs between groups. The sample size was not large enough to detect all types of, including less frequent or rare, AEs. The analyses of all immunogenicity endpoints were performed using randomized subjects who received CoVLP+AS03 or placebo from the Intent-to-Treat population set. Immunogenicity was evaluated by humoral immune response (NAb assays) and cell-mediated immune (CMI) response (ELISpot) induced in subjects on D0, 21 and 42. To assess the humoral immune response, the GMT was calculated and compared between CoVLP+AS03 and placebo groups using an ANOVA on the log-transformed titers. The log transformation was used to meet the normal assumption for the ANOVA. At each timepoint, the GMT and corresponding 95% CI of each treatment were obtained by exponential back-transformation of the least square mean. GMT were compared between study populations at D21 and D42 using an ANOVA. Comparison of D0 seronegative and seropositive values at D21 was conducted by unpaired t test of log-transformed values. Fisher’s exact test was used to compare seroconversion rate between the treatment groups. The 95% CI for seroconversion was calculated using the exact Clopper-Pearson method. The specific T helper type 1 (Th1) and Th2 CMI responses along with the corresponding 95% CI for the median induced on D0, D21 and D42 were measured by the number of cells expressing IFN-γ and IL-4 respectively, using ELISpot. The difference in IFN-γ and IL-4 response between treatment groups at each timepoint was compared using a non-parametric Wilcoxon Rank Sum Test. The difference in IFN-γ and IL-4 were also compared between study populations at D21 and D42 using a Kruskal-Wallis Test. Safety assessment are based on the Safety Analysis Set, i.e. all subjects who received CoVLP+AS03. Occurrence and incidence of safety events were reported for each treatment groups. No formal hypothesis-testing analysis of AE incidence rates was performed, and results were not corrected for multiple comparisons.

