## Supplemental Table 1 for "Phase 2 Randomized Trial of an AS03 Adjuvanted Plant-Based Virus-Like Particle Vaccine for Covid-19 in Healthy Adults, Older Adults and Adults with Comorbidities"

### SUPPLEMENTARY MATERIAL

**Supplemental Table 1: Summary of humoral responses in Healthy Adults, Older Adults, and Adults with Comorbidities**

| | Healthy Adults | | Older Adults | | Adults with Comorbidities | |
|---|---|---|---|---|---|---|
| | CoVLP 3.75 µg +AS03 | Placebo | CoVLP 3.75 µg +AS03 | Placebo | CoVLP 3.75 µg +AS03 | Placebo |
| **NAb Titer** | | | | | | |
| Day 0 | | | | | | |
| GMT (95CI) | 5.3 (5.1, 5.6) | 5.0 (NE, NE) | 5.2 (5.0, 5.5) | 5.5 (4.6, 6.6) | 8.9 (6.8, 11.6) | 13.0 (6.4, 26.2) |
| Day 21 | | | | | | |
| GMT (95CI) | 44.3 (35.8, 54.8) | 5.2 (4.8, 5.6) | 29.7 (23.7, 37.1) | 5.5 (4.6, 6.5) | 49.1 (31.8, 75.7) | 20.0 (8.2, 49.0) |
| Day 42 | | | | | | |
| GMT (95CI) | 2034 (1754, 2358) | 5.7 (4.7, 6.8) | 1918 (1571, 2342) | 5.6 (4.5, 6.9) | 1962 (1442, 2667) | 36.2 (12.5, 104.7) |
| **Seroconversion Rate** | | | | | | |
| Day 21 | | | | | | |
| SCR, % (95CI) | 51.3 (44.7, 57.8) | 0 (0.0, 8.2) | 38.4 (31.9, 45.1) | 0 (0.0, 7.9) | 44.2 (35.1, 53.5) | 7.7 (0.9, 25.1) |
| Day 42 | | | | | | |
| SCR, % (95CI) | 99.2 (97.0, 99.9) | 2.4 (0.1, 12.6) | 97.7 (94.7, 99.2) | 0 (0.0, 9.0) | 95.8 (90.4, 98.6) | 17.4 (5.0, 38.8) |

NAb: neutralizing antibody, GMT: geometric mean titer, 95CI: 95% confidence interval, SCR: seroconversion rate, NE: not estimable

1
